# Arousal network determines mean of systolic blood pressure and chemoreflex pathway determines dispersion of systolic blood pressure during N2 sleep in patients of moderate to severe obstructive sleep apnea

**DOI:** 10.64898/2026.01.12.25343259

**Authors:** Biswagourav Nahak, Dinu S Chandran, Karan Madan, Nasreen Akhtar

## Abstract

**Introduction:** Obstructive sleep apnea (OSA) is characterized by recurrent upper airway obstruction during sleep, leading to intermittent hypoxia, sleep fragmentation, and autonomic dysregulation. These disturbances contribute to nocturnal blood pressure (BP) surges and increased cardiovascular risk. While sleep-stage–dependent BP modulation is well established, high-resolution data on sleep-stage– specific systolic BP variability (BPV) in OSA are limited. This study examined beat-to-beat systolic BPV during N2 and rapid eye movement (REM) sleep and its relationship with sleep fragmentation indices in patients with moderate-to-severe OSA.

**Methods:** Clinically suspected OSA patients aged 18–65 years underwent overnight level I video polysomnography. Patients with apnea–hypopnea index (AHI) <15 events/h, central sleep apnea >35%, beta-blocker use, or excessive artifacts were excluded. Continuous systolic BP was estimated using a validated pulse transit time (PTT) method calibrated against cuff BP. Artifact-free 20-minute continuous segments were extracted separately from N2 and REM sleep. Sleep fragmentation metrics included number and duration of respiratory events, arousals, and area under the curve of oxygen saturation (AUC SpO₂). BPV indices included mean systolic BP, standard deviation (SD), and coefficient of variation (COV).

**Results:** Sixteen patients contributed 16 N2 and 16 REM segments. N2 sleep showed a higher number of respiratory events (p = 0.005) and arousals (p = 0.01) than REM sleep, while event duration and AUC SpO₂ were comparable. Mean systolic BP was 126 ± 12.5 mmHg during N2 and 130 ± 14.9 mmHg during REM, with REM significantly higher than N2 (mean difference −3.62 mmHg; p = 0.01). BP variability was highest during REM (SD 7.12 [4.91–9.25] mmHg; COV 5.95 [3.89–6.84%), intermediate during N2 (SD 5.25 [4.02–6.75] mmHg; COV 4.42 [3.17–4.83%), and lowest during wake (p < 0.001). In N2 sleep, arousal duration predicted mean systolic BP (R² = 0.48, p = 0.0025), while AUC SpO₂ strongly predicted SD and COV (R² = 0.74–0.79, p < 0.0001). REM-stage correlations were weaker and not predictive.

**Conclusion:** Systolic BP variability in OSA is strongly sleep-stage dependent, with REM sleep exhibiting exaggerated BP instability despite fewer respiratory events. Stage specific mechanisms linking arousals and hypoxia to BP regulation may underlie cardiovascular vulnerability in OSA.

## INTRODUCTION

Obstructive sleep apnea (OSA) is a sleep-disordered breathing condition marked by recurrent upper airway collapse during sleep, resulting in intermittent hypoxia, hypercapnia, and transient arousals (Dempsey et al., 2010).These episodes lead to sympathetic nervous system overactivation, cyclical oxygen desaturation, intrathoracic pressure swings, and fragmented sleep architecture (3). These disturbances contribute to alterations in cardiovascular homeostasis, manifesting as surges in Systolic Blood Pressure (SBP) and increased long-term risk for systemic hypertension and cardiovascular morbidity (4,5).

In healthy individuals, sleep is associated with a stage-dependent modulation of blood pressure. Non-rapid eye movement (NREM) sleep, particularly stages N2 and N3, is typically characterized by a reduction in mean systolic blood pressure (SBP) and enhanced baroreflex sensitivity, reflecting decreased sympathetic activity and increased parasympathetic tone (6,7). In contrast, rapid eye movement (REM) sleep is marked by irregular fluctuations in autonomic output, reduced baroreflex gain, and increased sympathetic bursts, especially during phasic REM, leading to elevated BP variability (8). The differential autonomic profile of these sleep stages underlines the importance of examining BP regulation in a sleep-stage–specific context.

The pathophysiological interplay between OSA and cardiovascular dysfunction is complex and multifactorial. Repeated apneas and hypopneas during sleep provoke hypoxemia and hypercapnia, which stimulate chemoreflex-driven sympathetic surges and lead to arousal-mediated blood pressure spikes. These transient hemodynamic events, when sustained nightly over long periods, contribute to endothelial dysfunction, increased arterial stiffness, and heightened cardiovascular risk (9,10). Of particular clinical interest, REM-predominant OSA has been associated with more pronounced cardiovascular instability and greater adverse cardiometabolic consequences than NREM-predominant OSA, owing to its unique autonomic profile and longer apnea durations (11,12).

Despite this knowledge, there remains a paucity of high-resolution, sleep-stage–specific data on beat-to-beat BP variability (BPV) in OSA patients. Traditional cuff-based methods are limited in their temporal resolution and fail to capture the transient BP surges associated with discrete apneic events or cortical arousals. The pulse transit time (PTT) method, a validated surrogate for continuous BP estimation, provides a non-invasive approach to evaluate dynamic BP changes at a beat-to-beat resolution throughout the sleep period (13,14).

Given that sleep in OSA is highly fragmented, with frequent transitions between stages due to arousals and respiratory events, averaging BP across the entire night may obscure important stage-specific variations. Therefore, analyzing BPV separately for N2 and REM sleep stages offers a more precise understanding of how OSA-related physiological stressors differentially affect cardiovascular regulation.

The hypothesis of this study is that sleep-stage–specific BPV, particularly during N2 and REM sleep, will be significantly influenced by indices of sleep fragmentation, including the frequency and duration of respiratory events, arousals, and the extent of oxygen desaturation. Specifically, we hypothesize that REM sleep, characterized by heightened sympathetic activity and diminished baroreflex sensitivity, will exhibit greater BPV and more pronounced hemodynamic responses to arousals and desaturation events compared to N2 sleep. By quantifying the associations between fragmentation indices and BPV using beat-to-beat PTT-derived systolic BP data, this study aims to delineate the mechanisms of cardiovascular instability in OSA and identify sleep stages that confer differential hemodynamic risk.

## METHODS

### Patients

Clinically suspected obstructive sleep apnoea patients between 18 to 65 years of age were screened using STOP-BANG Questionnaire from the sleep clinic of Department of Pulmonary Medicine and Critical Care and Sleep Disorders at the All-India Institute of Medical Sciences, New Delhi and from the community through personal or phone interviews and word of mouth. Written informed consent was obtained. Socio-demographic data and anthropometric measures including weight, height, body mass index, neck circumference, abdominal circumference, waist circumference, and waist-to-hip ratio were recorded. Participants were scheduled for an overnight level 1 video polysomnography at the Baldev Singh Sleep Electrophysiology Laboratory. Patients on beta blockers and those having central sleep apnea exceeding 35% were excluded. The study was conducted in accordance with the ethical principles of the Declaration of Helsinki. Ethical approval was obtained from the Institutional Ethics Committee, All India Institute of Medical Sciences (AIIMS), New Delhi (IEC No. _/), and written informed consent was obtained from all participants prior to enrollment.

### Polysomnography

All participants were instructed to come two hours before their normal sleep time to acclimatize to the laboratory environment. A single night of Level I attended video polysomnography using the SOMNOScreen Plus (SOMNOmedics GmbH™, Randersacker, Germany) was conducted, scored, and reported in accordance with the current international guidelines published by the American Academy of Sleep Medicine (version 2.3) (15). 10% of the studies were cross scored to ensure interrater reliability and consistency of scoring (16,17). Twenty-minute continuous segments were independently selected from N2 and REM sleep stages, primarily from the first half of the night. For each segment, the following metrics were extracted separately for N2 and REM: total number of respiratory events (apneas and hypopneas), total number of arousal events, total duration of respiratory events (in seconds), total duration of arousals (in seconds) and area under the curve of oxygen saturation (AUC SpO₂ in %mmHg). The area under the curve of oxygen saturation (AUC SpO₂) for each 20-minute segment was calculated by extracting pulse oximetry values at a sampling rate of 4 Hz. The average between two consecutive data points was computed, and the result was multiplied by the corresponding time interval to obtain the area in units of %·seconds. A higher AUC value indicates better oxygenation, reflecting less severe desaturation during the segment. So Sleep fragmentation was assessed using 20-minute continuous segments from N2 and REM sleep. Parameters included the number and total duration of respiratory events, number and total duration of arousals, and AUC SpO₂, providing a comprehensive measure of event frequency and associated physiological burden. The sleep fragmentation parameters were calculated for the 20-minute segments for each sleep stage in DOMINO^TM^ Software.

### Blood pressure measurement

Blood pressure (BP) measurement during the night was performed using a validated method involving pulse-transit-time (PTT) with the DOMINO software (v.2.2.0, SOMNOscreen-plus, Randersacker, Germany). The PTT method estimates blood pressure by measuring the time it takes for a pulse wave to travel between two arterial sites, typically from the heart to a peripheral site, such as the fingertip or earlobe. The computation of the systolic BP depends on a non-linear correlation between BP and PTT. It operates on the principle that as blood pressure rises, arteries become stiffer, which causes the pulse wave to travel faster and shortens the PTT. Conversely, a decrease in blood pressure results in a longer PTT. During sleep, this method allows for continuous, non-invasive estimation of blood pressure changes, making it useful for monitoring variations related to sleep stages, apneas, and arousals. For accurate measurement, PTT-based blood pressure estimation was initially calibrated against a standard cuff-based measurement for each patient at the beginning of the sleep study, as factors like heart rate variability, arterial compliance, and body position can influence the readings. The non-invasive measurement of BP using the SOMNOscreen plus has been validated following the European Society of Hypertension protocol (18,19). For assessment of blood pressure variability specific to a particular stage of sleep, twenty-minute segments of continuous, artifact free and raw RR interval data was exported in csv format to a commercial software Nevrokard BPV (Medistar Inc.). These signals were extracted from NREM and REM sleep and wake stages. This gave us 4,800 values of systolic BP for each 20-minute segment. The NREM segments were predominantly composed of stage N2 sleep (another contributor being N1), with N2 contributing more than 80% of each segment. Patients of moderate to severe OSA have very frequent respiratory events and arousals, which severely reduces the time they spend in N3. Hence, we have evaluated the associations only in N2 and REM stages of sleep. The wake blood pressure segments were extracted from the record of polysomnography after beginning of recording and before sleep onset. Patients with moderate to severe apnoea have frequent apneas and arousals, more than 15 per hour. Hence, extraction of artifact-free, apnoea free and arousal free segments of 20 minutes duration of the same sleep stage is difficult. The variability parameters of SBP were mean, Standard deviation (SD), Coefficient of variance (COV). The results were exported in excel format for analysis. We report only the above-mentioned variability parameters, not the time-domain analysis (SDSD, RMSSD) and frequency-domain analysis (LF, HF, VLF) of blood pressure as it requires data collected during stable physiological stages (20). Although time domain and frequency domain parameters of blood pressure would depict the physiological variations but we cannot use these values for analysis because OSA leads to frequent stage shifts and arousals, hence, it does not provide suitable conditions for accurate frequency-domain analysis of BP variability parameters (17).

### Statistical analysis

All data were entered in a database (GraphPad Prism Version 10.1.1 for Windows, GraphPad Software, Boston, Massachusetts USA, www.graphpad.com). A paired t-test or its non-parametric equivalent, the Wilcoxon test, was used to compare, arousal and respiratory event parameters between N2 and REM stages. For blood pressure variability parameters with a parametric distribution, repeated measures ANOVA followed by Tukey’s post hoc test for intra-group comparisons was employed to assess differences across wake, N2, and REM sleep stages. For non-parametric data, the Friedman test was used, followed by Dunn’s multiple comparison test to evaluate blood pressure variability across the same stages. An intra-group correlation matrix was obtained separately for N2 (NREM) and REM stages which correlated all the BPV and sleep fragmentation parameters, and simple linear regression was performed for variables with significant correlation. The level of statistical significance was set at 0.05. For correlation we have taken 0.5, r value as cutoff and for regression 0.3, R^2^ value as cutoff.

## RESULTS

Overnight polysomnography was performed for thirty patients. Out of thirty, seven patients were excluded for apnoea-hypopnea index less than 15 and three patients were excluded because artifacts in overnight polysomnography were more than 50%. Data was analysed for 20 patients. From the screening of 20 patient records, a total of 16 twenty-minute segments were obtained for each sleep stage (N2 and REM), resulting in 16 N2 and 16 REM segments for data analysis from 16 patients. Four patients were excluded due to the absence of continuous N2 sleep, defined as N2 comprising more than 80% of the selected segment duration. Demographic details and sleep phenotype of OSA patients who underwent overnight polysomnography are shown in table 1.

### Sleep fragmentation parameters in N2 and REM

There is a significant increase in the number of respiratory episodes (A+H) (p=0.005) and arousal events (p=0.01) in the N2 stage as compared to REM stage, whereas the duration of respiratory episodes, arousals, and oxygen saturation (AUC of SpO_2_) are comparable between N2 stage and REM stage as shown in Fig-1.

### Blood pressure variability parameters in Wake, N2 and REM

The mean of systolic BP during the wake, N2 and REM stages was 127 ± 11.63 mmHg, 126 ±12.49 mmHg and 130 ± 14.9 mmHg respectively. There was no statistically significant difference observed across stages (p=0.26), but in multiple comparisons of N2 and REM showed significant difference (p=0.01, mean difference of –3.62 (CI= −6.62 to –0.62)). Standard deviation of systolic BP is maximum in REM stage i.e. 7.12 (4.91-9.25) and is minimum during wake 3.92 (2.62-4.57), in N2 being 5.25 (4.02-6.75). Significant difference for SD was found across the stage (p=0.0001). Significantly higher values were found in REM compared to N2 (p=0.004) and REM compared to wake stage (p<0.0001). Coefficient of variation (COV) of systolic BP were significantly higher in the REM stage (5.95 (3.89-6.84) than in wake stage 3.13 (2.28-3.46) (p<0.0001), and higher in REM compared to N2 stage (4.42 (3.17-4.83, p=0.008) as shown in Fig-2.

### Correlation of sleep fragmentation with blood pressure variability

In the N2 sleep stage, arousal duration is positively correlated with mean (r = 0.69) of systolic BP. AUC SpO₂ has negative correlations with systolic BP SD (r = −0.51) and COV (r = −0.52), As shown in the figure 3. Arousal duration of the N2 stage significantly predicts mean of systolic BP (n=16, R^2^=0.48, p=0.0025, slope=0.042). Oxygen saturation predicts the standard deviation (n=16, R^2^=0.74, p<0.0001, slope = −0.00038) and COV (n=16, R^2^=0.79, p<0.0001, slope = −0.0003) of systolic BP significantly. as shown in the figure-3.

During the REM stage, Number of arousal events is positively correlated with mean SBP (r=0.5, p=0.04) and arousal duration is positively correlated with standard deviation of SBP (r=0.53, p=0.03), but number of arousal event and arousal duration not able to predict mean SBP (R^2^=0.25) and SD of SBP (R^2^=1.14) respectively in regression model.

## DISCUSSION

The aim of the present study was to investigate the relationship between sleep fragmentation characterized by apneic episodes, arousals, and oxygen desaturation, and sleep-stage specific systolic BP variability in patients with moderate to severe obstructive sleep apnea. Using high-resolution beat-to-beat systolic BP data at 4Hz synchronized with polysomnography, we delineated the hemodynamic impact of sleep architecture disturbances during N2 and REM stages of sleep.

### Sleep fragmentation parameters (Respiratory and arousal events in N2 vs REM)

In our study N2 sleep exhibited significantly higher frequencies of respiratory and arousal events compared to REM sleep, indicating increased sleep fragmentation. However, total respiratory event duration, total arousal duration, and oxygen desaturation burden AUC SpO_2_ were comparable between N2 and REM. This suggests that while N2 is more fragmented, the duration of these events remains similar across both sleep stages. Almeneessier et al. (11) reported no significant difference in the duration of obstructive respiratory events or the time spent with oxygen saturation below 90% when comparing REM and NREM sleep, indicating a similar degree of physiological impact across stages. In contrast, Han et al (21) observed that arousals were more frequent during NREM sleep than REM, suggesting greater sleep fragmentation during NREM despite comparable desaturation severity.

### Sleep Stage–Specific Blood Pressure Variability Patterns

Our findings indicate that mean of systolic BP differs significantly between N2 and REM and standard deviation of systolic BP significantly differ across N2 and REM stages. The mean of the systolic BP broadly reflects a regulatory set point around which the cardiovascular system dynamically operates on a beat-to-beat basis. It is an indirect indicator of the operational set point of background regulatory neural network. As the mean of systolic BP is changing across stages it suggests a repatterning in neural regulatory network across the stages of sleep in patients of OSA. These observations are consistent with prior reports that REM sleep, characterized by sympathetic surges, is associated with exaggerated cardiovascular instability in OSA (8,22). Loredo et al. (23) demonstrated elevated nocturnal BP surges during REM sleep in patients with OSA, independent of apnea index, suggesting heightened REM-specific blood pressure variability. REM sleep is a paradoxical state where increased sympathetic tone coexists with peripheral muscle atonia. During phasic REM, bursts of sympathetic activity driven by pontine-geniculate-occipital (PGO) waves coincide with irregular respiration, abrupt eye movements, and BP surges (Somers et al., 1993). This heightened sympathetic outflow, compounded by respiratory events in OSA, likely explains the elevated mean and variability of systolic BP observed in REM. Furthermore, baroreflex sensitivity is impaired during REM sleep (24) contributing to poor buffering of BP fluctuations.

Our regression models provide evidence that arousal duration during N2 are significant predictors of mean of SBP. This aligns with the work of Baguet et al. (2005), who reported that arousals, even more than respiratory events, drive nocturnal BP surges in OSA. Notably, the strong association of severity of oxygen desaturation with lower systolic BP variability indices (SD and COV) shows that an increase in desaturation severity increases dispersion in systolic BP values, as there is more fluctuation around a certain level of mean SBP. Thus, it appears that in N2 (NREM) sleep, the neural network responsible for arousals during sleep determine the operative level of the cardiovascular system like determining the mean of systolic BP but the chemoreflex driven circuit determines the dispersion pattern of the systolic BP.

In REM stage as the number of arousal events and arousal duration are positively correlated with mean and SD of SBP it shows the same arousal network might be operational during the REM stage but these arousal components are not able to predict the respective systolic BP Variability indices. One explanation is that the association is not strong enough for number and duration of arousals to drive mean and SD of systolic BP. Another explanation may be that the baseline sympathetic tone is already in overdrive during REM sleep and hence added perturbations in the form of arousals and desaturations do not alter the already increased entropy further. Factors such as OSA severity and age may modify the relationship. A bigger sample size would bring forth more robust associations. Further, the effect of respiratory component is not effective during REM, as the chemoreflex pathway is blunted due to under-responsive nucleus of the solitary tract (NTS) during REM. Somers et al have reported the maximum oxygen-triggered SBP value was significantly higher in rapid eye movement sleep than in non–rapid eye movement stage 1 sleep and non–rapid eye movement stage 2 sleep in patients with OSA (26). The temporal coupling of cortical arousal and sympathetic burst firing during REM may explain this effect (27). The degree of arousal was more influential in post-apneic BP elevation than the lowest oxygen saturation and respiratory event duration, where even brief arousals in REM cause disproportionately larger BP surges, likely due to lower vagal restraint and greater sympathetic readiness in this stage (28). From a pathophysiological perspective, our findings underscore the role of recurrent arousals and intermittent hypoxia in driving nocturnal BP instability in OSA, with distinct stage-specific manifestations. In normal scenario, without any sleep disrupting event, the regulation of arterial blood pressure (ABP) across sleep stages involves a complex interplay between reflex cardiovascular control and central autonomic commands, dynamically modulated by sleep-state-specific neural circuits. During NREM sleep, the baroreflex control is reset to lower levels of SNA and BP and to increased heart period. Such resetting would also contribute to the reduction in BP and SNA and the increase in heart period that occurs during NREMS compared with wakefulness (29,30). This autonomic stabilization is underpinned by heightened baroreflex sensitivity (baroreflex gain), which promotes blood pressure homeostasis by promptly counteracting fluctuations. In contrast, rapid eye movement (REM) sleep is characterized by a reorganization of sympathetic outflow, leading to variable changes in ABP—including phasic hypertensive surges and increased blood pressure variability (BPV).

While NREM sleep (particularly N2) is characterized by a high burden of respiratory events and arousals with relatively stable blood pressure, REM sleep exhibits significantly heightened BP variability including larger systolic excursions, increased standard deviation despite fewer respiratory disturbances. These findings can be explained with help of Figure 3.

In animal studies during NREM sleep, especially stage N2 (Figure 4, Panel A), the ventrolateral preoptic nucleus (VLPO) is tonically active, exerting GABAergic inhibition on wake-promoting and autonomic centres, including the rostral ventrolateral medulla (RVLM) and parabrachial nucleus (PBN) (31). This results in reduced sympathetic tone and stable vascular tone (31–34). Simultaneously, the nucleus of the solitary tract (NTS), a critical relay for baroreceptor input, remains functionally robust, maintaining baroreflex-mediated homeostasis (35) Despite a high density of arousals and respiratory events during N2, autonomic buffering remains intact, limiting the extent of BP variability. This explains our observation of high event frequency but low hemodynamic impact during NREM sleep, consistent with previous studies (8,36).

In contrast, REM sleep (Figure 3, Panel B) represents a neurophysiological state of autonomic disinhibition. During REM the neural circuitry in brainstem activates sublaterodorsal nucleus (SLD) and PPT which modulates both motor atonia and autonomic circuits (37,38). As shown in Figure 3, descending projection from the SLD and PPT to the RVLM may thus activate premotor neurons that increase SNA. neurons within the lateral PBN are most active during REMS, as compared with quiet wakefulness and NREMS (39), and these neurons may therefore contribute to resetting of the baroreflex and subsequent increase in SNA via their projection to the NTS (40,41). Compounding this, baroreflex sensitivity is suppressed, and the NTS becomes less effective in buffering BP fluctuations. This central configuration, combined with respiratory instability and cortical arousals, leads to abrupt, amplified BP surges—which we observed as significantly higher mean systolic BP and variability indices during REM.

Collectively, these animal experimental findings support the hypothesis that sleep-stage-specific neural circuits—particularly those involving the VLPO, PB, NTS, and RVLM—interact with cardiovascular reflexes to regulate Blood Pressure and its variability. Arousals, by disrupting this delicate balance, emerge as key contributors to transient cardiovascular instability, especially in pathophysiological conditions. Understanding these mechanistic underpinnings is crucial in elucidating the cardiovascular risks associated with sleep fragmentation and disorders like OSA.

Our study is limited by its small sample size and the exclusion of patients with AHI <15. Nonetheless, the focus on moderate-to-severe OSA ensures the validity of observed hemodynamic patterns. Future studies could explore stage-specific pharmacologic interventions or biofeedback-guided autonomic modulation targeting N2 arousals and REM sympathetic surges.

This study highlights the stage-specific contributions of sleep fragmentation and nocturnal hypoxia to systolic blood pressure variability in patients with obstructive sleep apnea (OSA). By identifying N2 sleep as a stage with a higher burden of respiratory and arousal events, and REM sleep as a phase of exaggerated blood pressure fluctuations, this research provides a nuanced understanding of cardiovascular stress in OSA. Such insights are clinically relevant because nocturnal blood pressure variability is a recognized risk factor for stroke, myocardial infarction, and cognitive decline, particularly in older individuals. Recognizing which sleep stages contribute to specific hemodynamic disturbances allows for more targeted therapeutic strategies—such as stage-specific ventilatory support, pharmacologic modulation of autonomic tone, or timing of antihypertensive medication. Ultimately, this approach could lead to personalized treatment paradigms aimed at reducing cardiovascular risk and improving long-term outcomes in patients with OSA.

## Data Availability

All data produced in the present study are available upon reasonable request to the authors

